# Making prognostic algorithms useful in shared decision-making: Patients and clinicians’ requirements for the Predict:Breast Cancer interface

**DOI:** 10.1101/2020.11.16.20232348

**Authors:** G.D. Farmer, G.M. Pearson, W.J. Skylark, A.L.J. Freeman, D.J. Spiegelhalter

## Abstract

**Objectives:** To develop a new interface for the widely used prognostic breast cancer tool: PREDICT. To facilitate shared decision-making around post-surgery breast cancer treatments. To derive insights into communicating the outputs of prognostic models to patients and their clinicians.

**Method:** We applied user-centred design principles in developing a new interface for PREDICT. The research involved online surveys, focus groups, meetings, and usability testing with patients, clinicians and the public.

**Results:** The new interface has been launched and delivers around 30,000 sessions per month. We identified several principles that are useful when communicating the output of prognostic models, including multiple presentation formats, and contextualising statistics. A programme of future work based on patient and clinician feedback has been developed, including the provision of quantitative data on the adverse effects of adjuvant breast cancer treatments.

**Conclusions:** For prognostic algorithms to fulfil their potential to assist with shared decision-making they need carefully designed interfaces. User-centred design puts patients and clinicians needs at the forefront, allowing them to derive the maximum benefit from prognostic models.

## 1 Introduction

Around 55,000 women are diagnosed with invasive breast cancer each year in the UK [1], with an estimated 2 million new cases each year worldwide [2]. These women, along with their healthcare professionals, need to make potentially life-altering decisions about their treatment. In order to inform those decisions, they need comprehensible and balanced information about the potential risks and benefits of the different treatment options [3].

In 2010 [4] a prognostic model called PREDICT was developed to estimate the survival benefits of different adjuvant (post-surgery) therapies for breast cancer. Individualised estimates are made on the basis of inputs describing the patient and their cancer. The PREDICT algorithm was embedded into a publicly available website, Predict:Breast Cancer, designed primarily for clinicians.

While the PREDICT statistical model has been extensively validated [6–11], the initial interface that allowed public access had not been designed or tested for comprehension and usability. It is well recognized that the ways in which numbers and evidence are presented can have a large impact on the audience’s perception of risks and benefits, and on decisions made as a result [12]. Careful design of the outputs of risk prediction algorithms is therefore necessary. Good design should improve comprehension, shared decision-making and standardization of treatment. We therefore set out to redesign the Predict:Breast Cancer interface, using the principles of user-centred design (UCD) [13]. This process resulted in a new interface collaboratively developed with patients and clinicians.

The user-centred design process is widely used in industry and considered best practice when designing interactive systems. The use of UCD in a medical context is less common but increasingly recognised as important [14–16]. We outline how we applied the UCD process to the Predict:Breast Cancer interface, and highlight insights that may help others ensure prognostic models maximise their potential to help with shared decision-making.

Post-redesign, the site is delivering over 30,000 sessions per month (see Figure 1). It is recommended by the National Institute for Health and Care Excellence (NICE) in the UK and endorsed by the American Joint Committee on Cancer [5].

**Figure 1.**
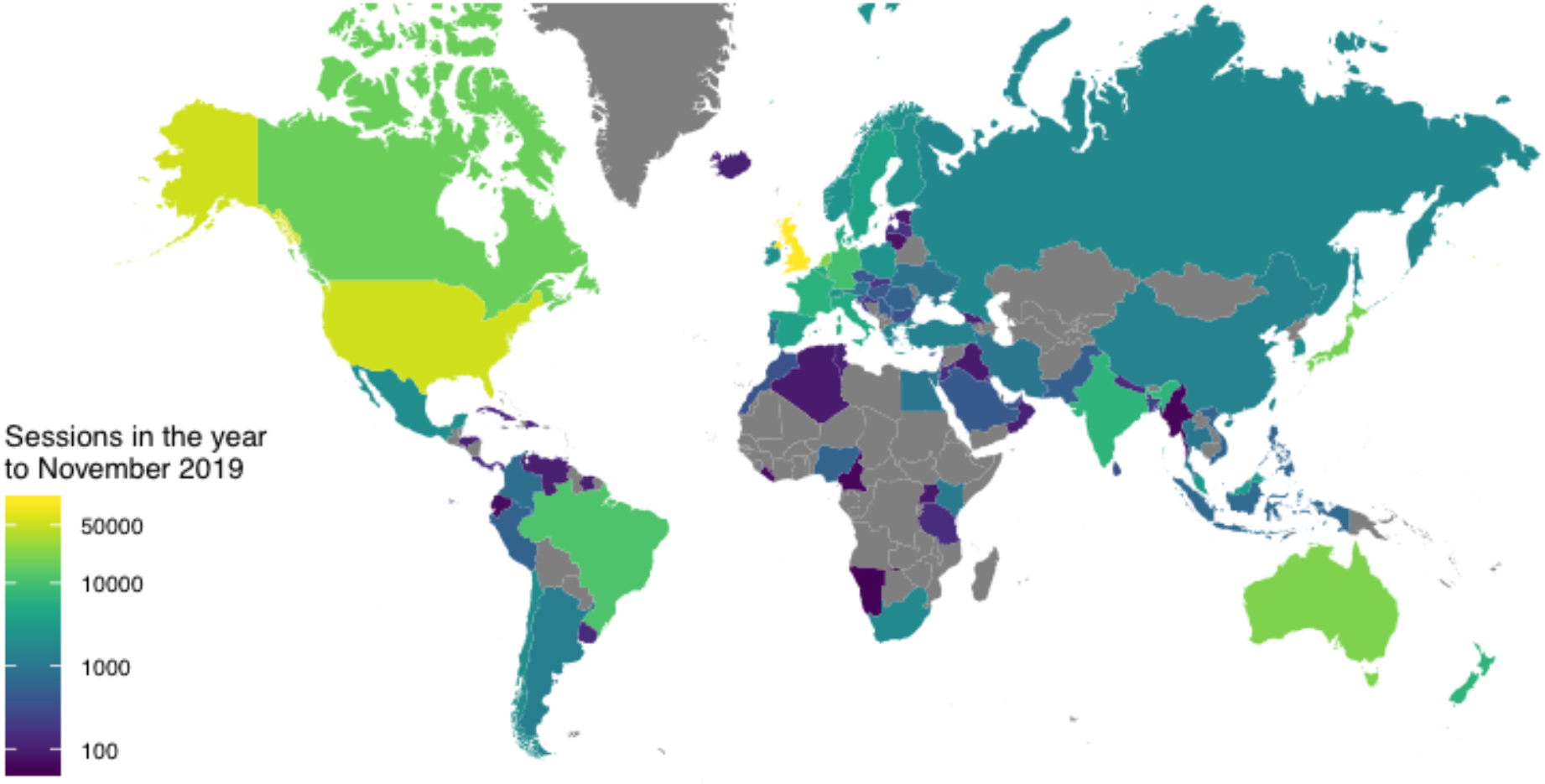
The top 100 countries by usage of the Predict:Breast Cancer website in the year to November 2019. The data are for ‘sessions’ as defined by Google Analytics.

## 2 Methods

This study was approved by the University of Cambridge Psychology Research Ethics Committee (PRE.2016.103). Participants gave informed consent before taking part.

### 2.1 Background research

The original interface allowed entry of parameters describing the patient and their cancer. The user then selected a combination of treatments and was presented with predicted survival at five and ten years. The display included a breakdown of how each treatment contributed to the estimated survival rate (see Figure 2).

**Figure 2.**
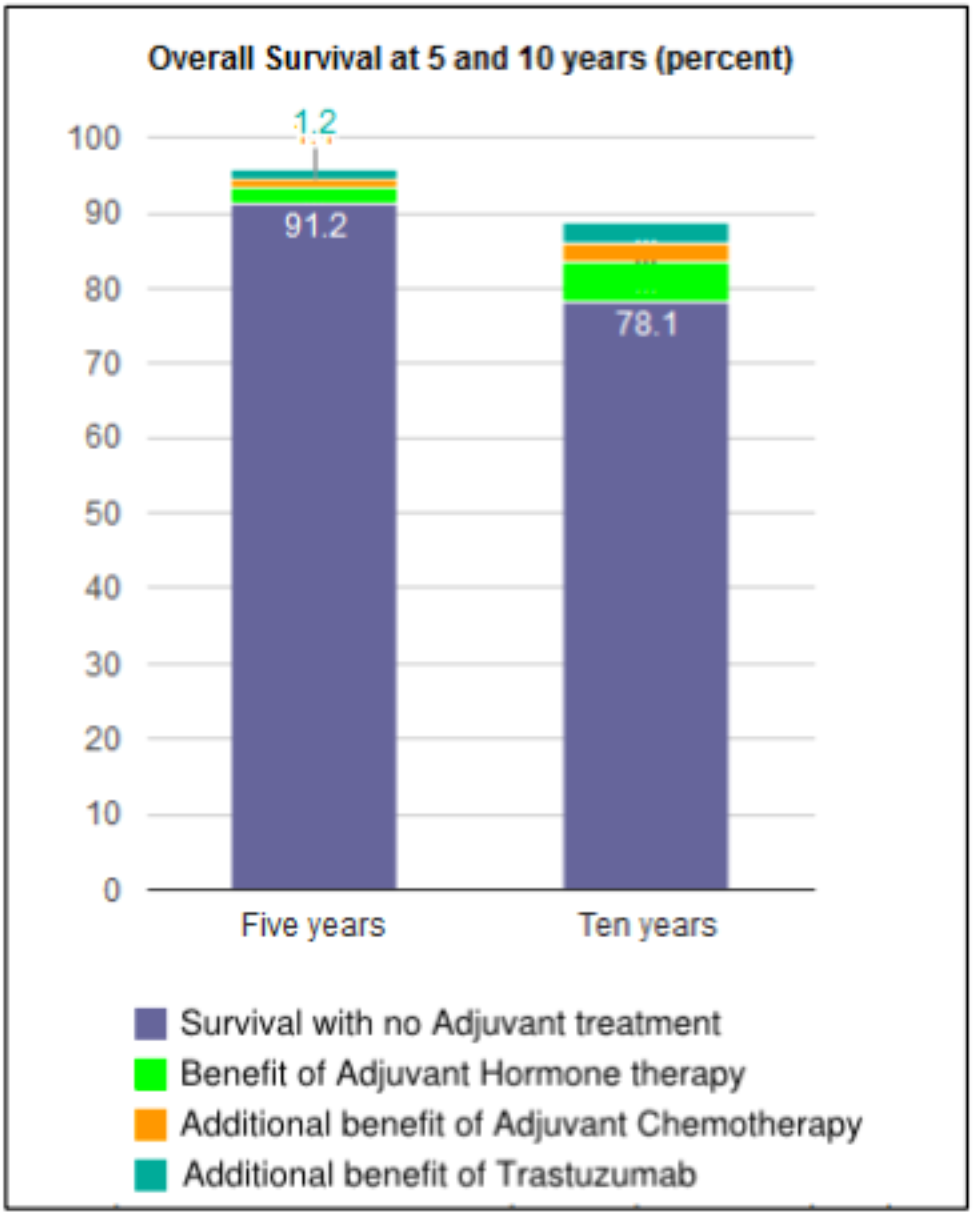
The original Predict:Breast Cancer interface. Hovering over a segment would reveal a pop-up display showing the increase in survival. The increases were also displayed as text below the chart.

To understand more about patient and clinician use, we searched online breast cancer forums for mentions of PREDICT, and observed a multi-disciplinary team meeting (MDT) at the Cambridge Breast Unit.

We conducted a focus group with women from the Cambridge area (*n*=7, Mean age = 46, *SD* = 13) in order to obtain more detailed feedback from naïve users. Participants were recruited via online community message boards. They took part in a session lasting 90 minutes that followed a semi-structured agenda. Participants discussed their expectations of a prognostic website, how comprehensible different visualisations were, and features not currently available that they would find desirable.

We also held a focus group with 18 breast cancer clinicians with different specialties at the Cambridge Breast Unit, addressing how they used the site, issues that they had with the current version, and their opinions on how it could be improved.

To validate and expand our feedback, we also carried out an online survey (*n* = 50, mean age = 37, *SD* = 10) with members of the public recruited from a national participant pool (prolific.co). Participants were given input parameters, asked to enter these into Predict:Breast Cancer, and then comment on the appearance, perceived trustworthiness of the site, and on how the results were displayed.

We also surveyed clinicians at the UK Breast Cancer Group (UKBCG) meeting in November 2016. Respondents (*n* = 75) were from 44 institutions across England, Scotland, Wales and Northern Ireland. The survey addressed the ways in which they were using Predict:Breast Cancer. Respondents were also asked to identify new inputs they would like to see in Predict:Breast Cancer and rated the desirability of potential new outputs; they also made comments in free-text fields.

### 2.2 Iterative Development

We developed prototypes informed by the background research. Wireframes^1^ were used in order to encourage feedback that included wholesale changes to the design. Later iterations with increased functionality and realism resulted in incremental feedback until a final prototype was ready for launch.

We obtained feedback and suggestions for improvement from breast cancer clinicians at Addenbrooke’s and Hillingdon hospitals. Clinicians were shown the wireframes and given a demonstration of example interactions that it would allow (e.g., selecting different timeframes over which to see the predictions of the algorithm). Participants were then invited to discuss problems, benefits and alternative solutions.

After the early prototype feedback was addressed, a functioning prototype was made available to participants for usability evaluation. These included breast cancer clinicians (*n*=7) who had indicated they were willing to be contacted when we surveyed the UK Breast Cancer Group (UKBCG) 2016 conference, and breast cancer patients (*n* = 12, mean age = 52, *SD* = 10) recruited from a pool of participants who had previously worked with our centre. Participants were asked to interact with the website whilst software recorded video, audio and mouse movements. Each participant undertook a pre-set task of entering data and interpreting the results, as well as some unstructured interaction in which they were asked to use the site as they would normally.

In the final stage we employed a graphic designer to develop a professional-standard design for the website based on the participant feedback. This prototype was made available to the patient advocacy group Independent Cancer Patients’ Voice, who reviewed the site before the final iteration.

## 3 Results

### 3.1 Background research

We analysed comments from our online survey, patient forums, and the public focus group to derive several themes that characterised patients’ and members of the public’s reaction to using the Predict:Breast Cancer website.

#### Fear of a poor prognosis

Many participants indicated that they would be put off using a prognostic tool for fear of receiving worse news than they expected. For some, there was a sufficiently strong sense of foreboding that using the tool would simply be too frightening (e.g., Box 1 comment 1).

**Box 1.**
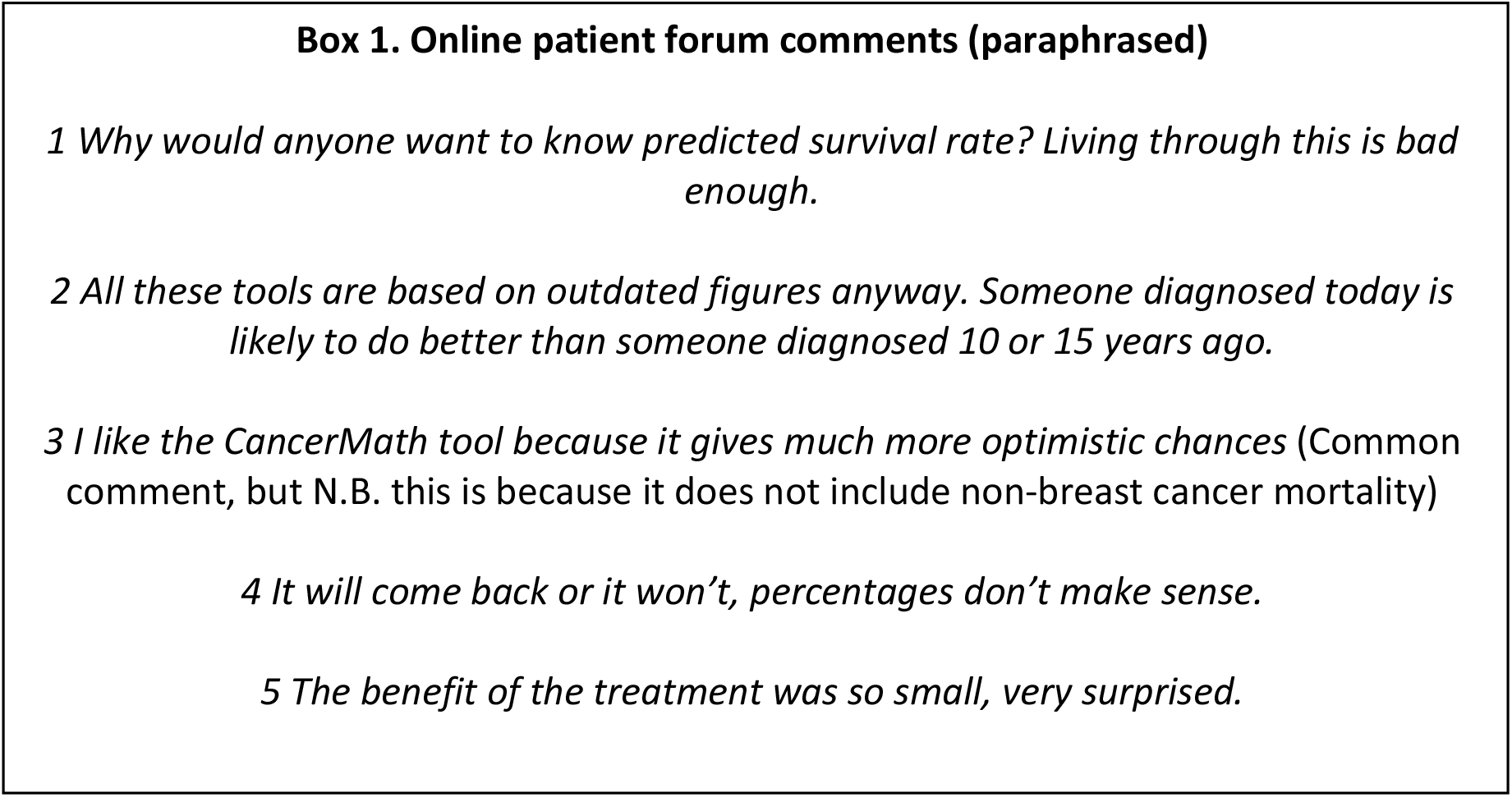
Illustrative comments about the Predict:Breast Cancer tool on patient forums.

#### Appearance and trustworthiness

Participants in the online survey and focus group identified that the site appeared ‘clinical’, ‘cold’ and ‘basic’. The clinical tone resulted in a perceived lack of empathy for the user while the basic style risked a lack of authority. Focus group participants also indicated that while icon arrays were a good communication device in general, the use of human-shaped icons would be too upsetting, and that abstract representations were preferable. The site was perceived to be trustworthy though this was in large part because of its affiliation to the UK’s National Health Service.

#### Scepticism about statistics and predictions

A common response was to question the accuracy of the predictions. This scepticism was a result of the perception that the tool did not ask for all the possible relevant information (e.g., exercise levels or type of surgery). Participants felt therefore that where their individual circumstances were not taken into account by the tool, this limited its ability to provide a useful prediction. A related response was the perception that percentages felt meaningless for an individual. Some participants felt that applying a percentage chance to a categorical event (survival) was hard to understand (e.g., Box 1 comment 4). Another important factor was that other tools appear to provide different estimates. This potentially has the effect of reducing trust in all tools, or leading patients to use those tools that they perceive to provide more optimistic outlooks (e.g., Box 1 comment 3)^2^. Finally, participants felt that the predictions must be based on old data, and therefore that they would not be up to date and include recent developments in treatment (e.g., Box 1 comment 2).

#### Information on side-effects

Participants identified that the tool only supplied quantitative information about the benefits of adjuvant therapies, and that they would also want to know about the side-effects and their likelihood. Participants felt that in some cases side-effects might be severe enough that they would refuse a treatment with only a small benefit. Of particular note was that some patients in the forum indicated surprise at how little potential benefit they were receiving from their treatment regimens (e.g., Box 1 comment 5).

#### Interface

Several aspects of the existing interface were problematic for participants. The primary concern was that small increments were difficult to read on the bar chart (see Figure 2). Many participants in the online survey also stated that the wording ‘an extra x women would survive with [treatment] confusing and would prefer the total number surviving instead.

Analysis of the UKBCG survey, observation of a breast MDT and comments from the clinician focus group revealed themes focussed more on the usability and functionality of the site:

#### Simplicity

Clinicians were very keen that the tool should be quick to use and with minimal constraints on access. This included highlighting technical restrictions in hospitals such as poor WiFi coverage and outdated browsers. Feedback also emphasised that the site should not require users to log in or demand inputs that might not be available. ‘Keep it simple’ was a common comment, and simplicity was a much-appreciated feature of the original interface.

#### Additional functionality

Many clinicians were keen to see new treatments added to the prognostic model (e.g., radiotherapy and bisphosphonates). Clinicians also highlighted additional outputs that would be useful such as different time frames for the model’s predictions. Clinicians also reported that not all patients would have access to computers and internet, meaning the ability to print the outputs was particularly useful.

#### Different use cases

Observation of the MDT and feedback from clinicians highlighted the variety of contexts in which the tool is used. In MDTs the tool was most often used in cases whether it was unclear if a sufficiently large increase in survival would be obtained. Consequently, the ability to inspect small increments in the tool’s output was critical. Clinicians also used the tool with patients during consultations to help explain the benefits of treatments. Additional uses identified from the survey included research, teaching and meeting preparation.

### 3.2 Interface Development

The new Predict:Breast Cancer website can be visited at the following address: https://breast.predict.nhs.uk/ and the open-source code is available here: https://github.com/WintonCentre/predict-v21-main

#### New Features

The finished interface includes many new features derived from the background research and feedback during iterative development process. These include: The option to choose a preferred method for displaying the results (Fig 3a); The inclusion of non-breast cancer mortality (Fig 3b); clickable information icons which display explanations of inputs and outputs (Fig 3c); The ability to specify the time range over which to display the predictions (Fig 3d); The optional provision of precision estimates around the predictions (Fig 3e);

**Figure 3.**
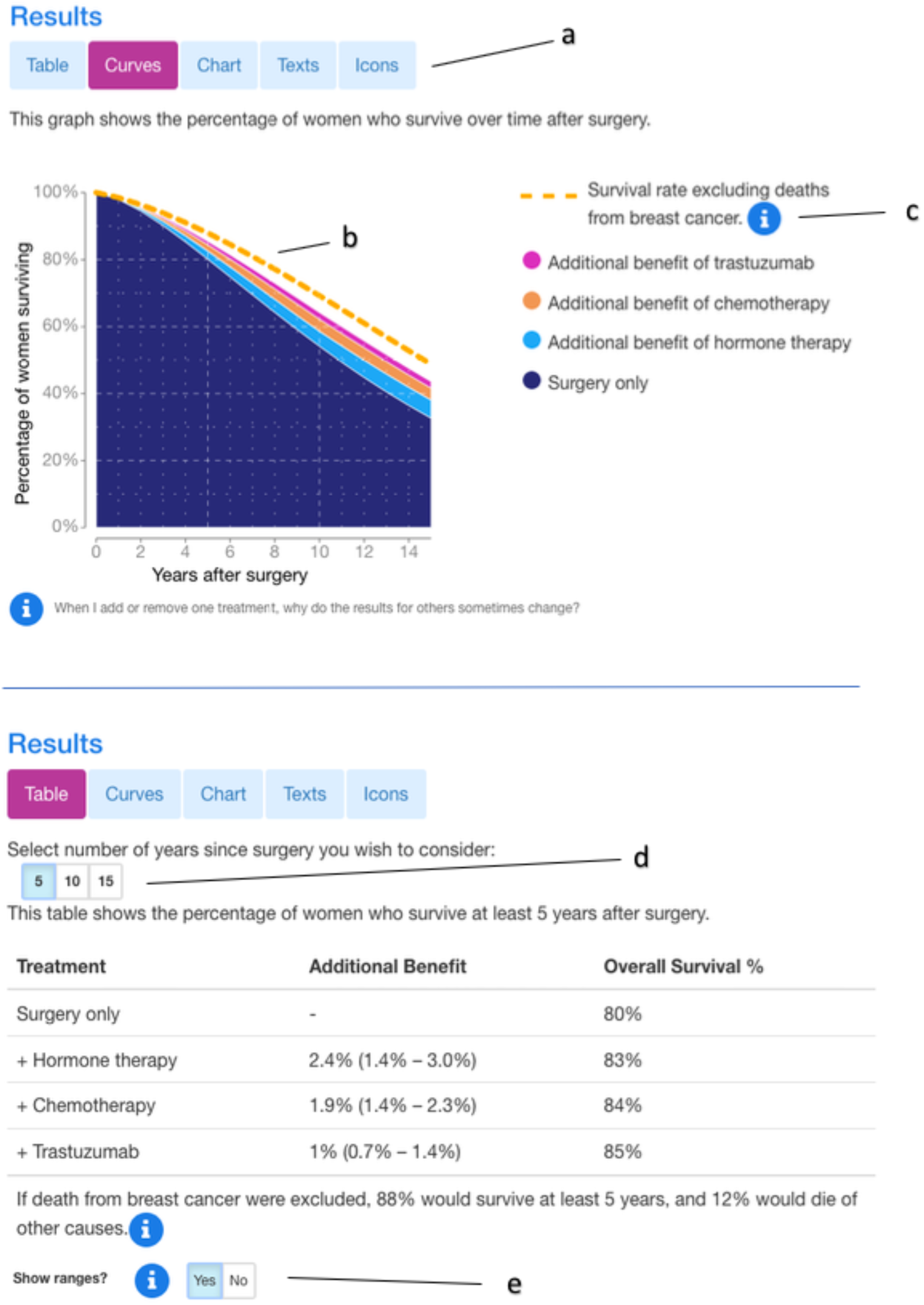
Display options for the new Predict:Breast Cancer website showing a survival curve in the top panel and a tabular format in the bottom panel. Users can choose from these formats and three others. a) choice of display b) survival rate excluding breast cancer c) information icons d) choice of timeframe e) optional prediction ranges.

The research and design process also revealed additional inputs and treatments that patients and clinicians would like to see as part of the model. Data was available for some of these, consequently bisphosphonates and extended hormone therapy have been added as treatments, while menopausal status has been added as an input. The Predict:Breast Cancer site now also has updated FAQs and technical information pages for non-experts. The site has been translated into Spanish and a graphic designer was employed to create a coherent professional appearance for the site.

#### Future features

The design process allowed us to identify additional inputs and outputs that clinicians and their patients would find useful. These data provide a valuable insight into how users would like the site to evolve and they form the basis for a plan of future research and development. Crucially, these development plans are user-driven and reflect how patients and clinicians would like the algorithm and interface to work in order to be maximally useful.

One requested function was the ability to use the tool for estimating the impact on survival of *stopping* adjuvant treatments. Patients may request this information because the side-effects of treatments are often unpleasant enough that they want to re-assess whether the treatments are worth continuing with.

A common query among patients was how to gauge the likelihood and severity of side-effects. While qualitative information on side-effects is available, there is very little quantitative information. Work is now underway to provide these data.

A related issue is the increase in mortality that some treatments cause. This can, in some patients, outweigh any benefit, meaning the treatment would result in a drop in predicted survival rates. This vitally important component is not currently included in Predict:Breast Cancer, and research is underway to determine how best to display such a ‘negative benefit’ unambiguously in each of the graphical formats used in Predict:Breast Cancer.

Two frequently requested features were co-morbidities as a patient input, and radiotherapy as a treatment option. The data are not currently available to model the effects of co-morbidities. However, radiotherapy as a treatment option is being modelled and will be added to a future release of the site.

Finally, both clinicians and patients expressed the desire to see the chances of breast cancer recurrence as well as survival. This can be modelled, and a programme of work is underway on how best to visualise this.

### 3.3 Insights on communicating prognostic predictions

In this section we relate the interface design choices to the background research and derive some broad themes to consider in the development of similar tools. The changes made to the Predict:Breast Cancer interface can be thought of as falling under three broad themes: Flexible and responsive interaction, contextualising statistics, and addressing gaps in knowledge.

#### Flexible and responsive interaction

It is important for an interface to allow the user to interact with the model’s predictions in a way that suits their needs. Rather than try and develop a ‘best’ way of representing the information, it can be useful to offer many options thereby maximising the number of users and use-cases that can be catered for. In the new interface, users have the ability to choose different ways of visualising the predictions. This allows clinicians to inspect tables for rapid and precise access to small increments, while survival curves allow patients to see how treatment decisions now impact on survival rates over time. Other benefits include catering to a user’s individual preferences when it comes to interpreting data and allowing users to check their understanding against a different presentation format. The new displays also update immediately upon a change to one of the input parameters. This responsivity allows for rapid comparison of different treatments through seeing the displays update instantaneously as treatments are added or removed. The design process revealed that both patients and clinicians can increase their understanding by seeing how changes to input parameters change predictions. Finally, allowing the user to choose the timeframe they want to visualise allows clinicians to select a period that is suitable given a patient’s age and prognosis.

#### Contextualising statistics

It is possible to communicate numbers perfectly accurately but leave the recipient none the wiser. The provision of contextual information can be critical in turning a number into information. The addition of non-breast-cancer deaths provides essential context for a survival probability. A survival estimate of 60% will seem less severe when it is in the context of a survival probability of 70% for similar women without breast cancer. This also makes clear the potential range for the treatment benefit – in this example, 10% would be the maximum possible increase in survival. Other information, such as uncertainty estimates, allowed the tool to convey not just the point estimate for survival (e.g., 60%) but also a range indicating the likely variability around the estimate, (e.g., 57% to 63%), especially important to take into account when the potential benefits are small.

#### Address gaps in knowledge

Most users of an interface, whether an expert clinician or new patient, will have questions about how it works. It is important to address these explicitly. One issue we identified was that some clinicians did not use PREDICT because the displayed benefit from the drug trastuzumab was less than their expectation based on clinical trials^3^. The interface now explicitly addresses this issue with the aim of increasing comprehension, trust and usage among clinicians. For patients, supporting information is important in several respects. The information icons provide help on how to complete the inputs accurately and on interpreting the outputs. This is not simply a usability issue, it increases a user’s confidence in the results, and helps them digest information outside of consultations with clinicians. We also sought to address users’ lack of confidence in statistical predictions by adding FAQ and technical information sections to the website. These explicitly address concerns raised in the background research and development process.

## 4 Discussion

The new Predict:Breast Cancer website is live and delivering thousands of sessions worldwide per week. Its development was based on a combination of broad background research and user-testing with the target audiences. By involving users in the design process, we believe it has enhanced the potential for shared decision-making and informed consent. Furthermore, the interface has now been successfully applied to a similar prognostic algorithm for prostate cancer (https://prostate.predict.nhs.uk) where its effects on decision-making and patient satisfaction are being evaluated in clinic [17,18].

We have also highlighted some general principles that will serve as useful guidelines for developers of other prognostic models and interfaces. These cannot substitute for engaging with users in each case, but we hope they serve as a framework for generating designs, especially where resources are not available for extensive consultation with users.

It is particularly worth highlighting that a large majority of participants showed a strong appetite for the information contained in Predict:Breast Cancer. It was also striking that patients wanted to trade-off the benefits of treatments with their potential adverse effects, in order to make a more balanced assessment. It may be that patients would prefer not to have treatment with a small average reduction in mortality if it means substantially lowering quality of life. For those who take treatment, awareness of the likelihood and severity of potential side-effects (and ways to ameliorate them) could help increase adherence to the treatment and reduce worry about symptoms that arise. This is a critical feature that is currently lacking in Predict:Breast Cancer.

During the research we found that patients would like to use Predict:Breast Cancer to understand the consequences of stopping treatments when they are suffering with side-effects. Analysis of patient forums also indicated some patients’ surprise at how little benefit they were receiving from their treatment. These all point to the importance of providing full information, including the potential harms of treatments.

### 4.1 Study limitations

To ensure the effectiveness of design decisions there is no substitute for feedback from prospective users at all stages of development. The inherent weakness in this approach is also its principal strength: the end product is bespoke, meaning that it won’t generate hard and fast rules for communication but will maximise efficiency in communicating with the particular audience being targeted. Fortunately, the method itself is generalisable and relatively simple to implement.

We did not recruit women who were recently post-surgery and currently making decisions about adjuvant treatments. It was decided that this was an inappropriate time to intervene in these patients’ treatment, and we instead sought inputs from clinicians, and patients who had previously been through the process.

### 4.2 Clinical implications

Epidemiological models can provide vital information for clinicians and patients deciding on which treatments to take. This in turn can lead to increased survival rates and all-round improved decision-making. User-centred design can remove the barriers to these benefits and, by engaging with patients, set an agenda for future development that really addresses their needs. In the case of Predict:Breast Cancer this has led to the first quantitative review of side-effects for many adjuvant therapies and additional modelling to address the requested additional treatments and outputs.

### 4.3 Conclusions

Prognostic algorithms have great potential for improving shared decision-making and informed consent. If made available in clinic, they can ensure that patients and healthcare professionals receive accurate and relevant information about the personalised benefits and harms of different treatment options. In order to make such complex and emotionally difficult information available in a clear, unambiguous, and sensitive manner, user-centred design is critical. Algorithms need to be statistically validated, but must also be easy to use, trustworthy, and produce outputs that are clear and useful to their users.

## Data Availability

The data that support the findings of this study are available from the corresponding author upon reasonable request.

## Acknowledgments

This work was supported by the David and Claudia Harding Foundation. GF was also supported by a Wellcome ISSF award (204796/Z/16/Z).We are very grateful to the patients and clinicians who volunteered to take. Paul Pharoah initiated the collaboration to develop a new interface and provided comments on a draft of this manuscript. Zsófia Szlamka and Leila Finikarides helped with data collection and participant recruitment. We are grateful to Independent Cancer Patients’ Voice for providing expert feedback. Graphic design was by Alison Norden.

## Footnotes

1 A sketched interface used to communicate potential functions and layouts

2 Major discrepancies were due to some tools excluding mortality not related to breast cancer.

3 Because the displayed increase in survival a treatment can provide depends on whether other treatments previously applied in the algorithm have already taken up some of the total possible increase.

## References

1 Cancer Research UK. https://www.cancerresearchuk.org/health-professional/cancer-statistics/statistics-by-cancer-type/breast-cancer (accessed 8 May 2019).

2 Bray F, Ferlay J, Soerjomataram I, et al. Global cancer statistics 2018: GLOBOCAN estimates of incidence and mortality worldwide for 36 cancers in 185 countries. CA Cancer J Clin 2018;68:394–424. doi:10.3322/caac.21492

3 Neuberger, Hale, Kerr, et al. Montgomery (Appellant) v Lanarkshire Health Board (Respondent) (Scotland). 2015.

4 Wishart GC, Azzato EM, Greenberg DC, et al. PREDICT: a new UK prognostic model that predicts survival following surgery for invasive breast cancer. Breast Cancer Res 2010;12:R1. doi:10.1186/bcr2480

5 Kattan MW, Hess KR, Amin MB, et al. American Joint Committee on Cancer acceptance criteria for inclusion of risk models for individualized prognosis in the practice of precision medicine. CA Cancer J Clin 2016;66:370–374.

6 Candido dos Reis FJ, Wishart GC, Dicks EM, et al. An updated PREDICT breast cancer prognostication and treatment benefit prediction model with independent validation. Breast Cancer Res 2017;19:1–13. doi:10.1186/s13058-017-0852-3

7 Engelhardt EG, van den Broek AJ, Linn SC, et al. Accuracy of the online prognostication tools PREDICT and Adjuvant! for early-stage breast cancer patients younger than 50 years. Eur J Cancer 2017;78:37–44. doi:10.1016/j.ejca.2017.03.015

8 Wong HS, Subramaniam S, Alias Z, et al. The predictive accuracy of PREDICT: A personalized decision-making tool for southeast Asian women with breast cancer. Med U S 2015;94:e593. doi:10.1097/MD.0000000000000593

9 Maishman T, Copson E, Stanton L, et al. An evaluation of the prognostic model PREDICT using the POSH cohort of women aged ≤40 years at breast cancer diagnosis. Br J Cancer 2015;112:983–91. doi:10.1038/bjc.2015.57

10 Wishart GC, Rakha E, Green A, et al. Inclusion of KI67 significantly improves performance of the PREDICT prognostication and prediction model for early breast cancer. BMC Cancer 2014;14:1–6. doi:10.1186/1471-2407-14-908

11 Wishart GC, Bajdik CD, Dicks E, et al. PREDICT Plus: Development and validation of a prognostic model for early breast cancer that includes HER2. Br J Cancer 2012;107:800–7. doi:10.1038/bjc.2012.338

12 Trevena LJ, Zikmund-Fisher BJ, Edwards A, et al. Presenting quantitative information about decision outcomes: a risk communication primer for patient decision aid developers. BMC Med Inform Decis Mak 2013;13:S7. doi:10.1186/1472-6947-13-S2-S7

13 Norman DA, Draper SW. User centered system design: New perspectives on human-computer interaction. CRC Press 1986.

14 Agoritsas T, Heen AF, Brandt L, et al. Decision aids that really promote shared decision making: the pace quickens. BMJ 2015;350. doi:10.1136/bmj.g7624

15 Montori VM, Breslin M, Maleska M, et al. Creating a Conversation: Insights from the Development of a Decision Aid. PLoS Med 2007;4. doi:10.1371/journal.pmed.0040233

16 Farmer, G.D., Gray, H., Chandratillake, G. et al. Recommendations for designing genetic test reports to be understood by patients and non-specialists. Eur J Hum Genet (2020).

17 Thurtle DR, Greenberg DC, Lee LS, et al. Individual prognosis at diagnosis in nonmetastatic prostate cancer: Development and external validation of the PREDICT Prostate multivariable model. PLoS Med 2019;16.

18 Thurtle DR, Jenkins V, Pharoah PD, et al. Understanding of prognosis in non-metastatic prostate cancer: a randomised comparative study of clinician estimates measured against the PREDICT prostate prognostic model. Br J Cancer 2019;121:715–8. doi:10.1038/s41416-019-0569-4

